# Dynamics of anti-SARS-CoV-2 seroconversion in individual patients and at the population level

**DOI:** 10.1101/2022.03.20.22272651

**Authors:** Alina Szewczyk-Dąbrowska, Wiktoria Budziar, Krzysztof Baniecki, Aleksandra Pikies, Marek Harhala, Natalia Jędruchniewicz, Zuzanna Kaźmierczak, Katarzyna Gembara, Tomasz Klimek, Wojciech Witkiewicz, Artur Nahorecki, Kamil Barczyk, Urszula Grata-Borkowska, Krystyna Dąbrowska

## Abstract

The immune response and specific antibody production in COVID-19 are among the key factors that determine both prognostics for individual patients and the global perspective for controlling the pandemics. So called “dark figure”, that is, a part of population that has been infected but not registered by the health care system, make it difficult to estimate herd immunity and to predict pandemic trajectories.

Here we present a follow up study of population screening for hidden herd immunity to SARS-CoV-2 in individuals who had never been positively diagnosed against SARS-CoV-2; the first screening was in May 2021, and the follow up in December 2021. We found that specific antibodies targeting SARS-CoV-2 detected in May as the “dark figure” cannot be considered important 7 months later due to their significant drop. On the other hand, among participants who at the first screening were negative for anti-SARS-CoV-2 IgG, and who have never been diagnosed for SARS-CoV-2 infection nor vaccinated, 26% were found positive for anti-SARS-CoV-2 IgG. This can be attributed to of the “dark figure” of the recent, fourth wave of the pandemic that occurred in Poland shortly before the study in December. Participants who were vaccinated between May and December demonstrated however higher levels of antibodies, than those who undergone mild or asymptomatic (thus unregistered) infection. Only 7% of these vaccinated participants demonstrated antibodies that resulted from infection (anti-NCP). The highest levels of protection were observed in the group that had been infected with SARS-CoV-2 before May 2021 and also fully vaccinated between May and December.

These observations demonstrate that the hidden fraction of herd immunity is considerable, however its potential to suppress the pandemics is limited, highlighting the key role of vaccinations.

## 1. Introduction

The new virus SARS-CoV-2 identified at the end of 2019, through its rapid spread, has caused a global epidemic, overloading or even paralyzing healthcare systems worldwide [1]. To date, efforts directed at therapeutic options for coronavirus disease remain limited, which is why it is so important to target the fight against the pathogen by understanding COVID-19 immunology, which should translate into reducing the spread of SARS-CoV-2 and ending the pandemic [2,3].

Despite over 440 million confirmed cases of COVID-19, the true prevalence of infection remains significantly underestimated due to a fraction of asymptomatic and oligosymptomatic infections as well as to limited abilities of robust diagnostics worldwide [4,5,6,7]. The serological response and the association with clinical symptoms are important in understanding the pathogenesis of COVID-19 [8]. The kinetics of antibody responses after COVID-19 are increasingly described in the literature to date, with early data showing divergent results regarding antibody responses in groups differing in disease severity. Most of the immunological studies, however, show that severe COVID-19 triggers an earlier and more intense immune response in hospitalized patients [8,9,10]. Many studies focus on assessing the immune response after vaccination, typically showing higher antibody titres in convalescents than in healthy individuals. The results of cohort studies that are currently available confirm the protective effect of natural infection. This protection persists for five to eight months, while pharmaceutical companies have defined the expected duration of the immune response of the vaccine as six months after the second dose [11]. Also, a significant decrease in antibody levels in COVID-19-patients more than 6 month after being discharged from the hospital has been observed [12].

Due to the rapid spread of the epidemic, there is still insufficient knowledge about safe and effective vaccination strategies, and it is not known which vaccination strategies will be most beneficial [11]. More complex is the question of how far observations from infections with one SARS-CoV-2 variant can be extrapolated to a different one. In view of the above, studies on antibody titres and the duration of the immune response in different groups of individuals appear to be necessary to understand and predict the protective mechanisms of anti-SARS-CoV-2 antibodies in COVID-19.

Here we present a follow-up study for a population screening for SARS-CoV-2 specific antibodies in Polish citizens (N=501) who had never been positively diagnosed with or vaccinated against SARS-CoV-2 [7]. In that group we identified a significant, hidden fraction of individuals who had developed anti-SARS-CoV-2 antibodies. In this study we evaluated IgG specific to viral nucleocapsid protein (NCP) and to receptor binding domain within spike protein (RBD) 7 months after the first screening for SARS-CoV-2 antibodies, seeking to understand how the antibody levels change with time in patients who were affected by an asymptomatic to a mild course of infection (non-hospitalized). We include an additional immunological factor, vaccination, since a part of that group was vaccinated during 7 months after the first screening for SARS-CoV-2 antibodies. We also include observation of kinetics on the same types of antibodies in patients hospitalized due to COVID-19.

## 2. Material and methods

### Population screening (inclusion/exclusion criteria)

This study is a follow-up of the anti-SARS-CoV-2 antibody screening completed on 15 and 22 May 2021 in healthy individuals who had not been previously vaccinated against COVID-19 and had never been positively diagnosed for SARS-CoV-2 infection [7]. In the period between May and December 2021 the fourth wave of COVID-19 occurred, with the Delta variant contribution assessed as almost 100% from mid-October until the end of the year. That wave peaked around 28th November to 10th December, with 7-day averages exceeding 23 000 confirmed SARS-CoV-2 infections [13].

The blood samples for re-evaluation of the anti-SARS-CoV-2 antibodies were collected in Wroclaw (Poland) on December 11, 2021, that is approximately 7 months after the first testing. Out of 501 people screened for SARS-CoV-2 antibodies in May 2021, 109 people were included in the follow-up study, considering previous serological status as identified in the first study, status of vaccination, their availability and agreement to participate in the repeated testing. Information on participants’ comorbidities and on their occupation as potentially linked to frequency of social interactions was available from data completed at the first screening on May 2021. All participants were interviewed for possible COVID-19 diagnosis, for current infection symptoms, as well as smoking status, their height and weight. Participants who were affected by diagnosed COVID-19 between May and December 2021 were excluded. Participants who were partially vaccinated or fully vaccinated but less than 4 weeks before December 11, 2021 were excluded; two doses of Pfizer-BioNTech or Moderna vaccine, or a single dose with the Johnson & Johnson vaccine were considered the full vaccination.

The following groups were selected:

#### Group 1 (N-, V-)

Participants seronegative for SARS-CoV-2 antibodies in May 2021 (anti-NCP-negative), non-vaccinated; N=38

#### Group 2 (N-, V+)

Participants seronegative for SARS-CoV-2 antibodies in May 2021 (anti-NCP-negative), fully vaccinated before December 2021; N=29

#### Group 3 (N+, V-)

Participants seropositive for SARS-CoV-2-specific IgG antibodies in May 2021 (anti-NCP-IgG positive) and non-vaccinated; N=25

#### Group 4 (N+, V+)

Participants seropositive for SARS-CoV-2-specific IgG antibodies in May 2021 (anti-NCP-IgG positive), fully vaccinated before December 2021; N=17

Of note, all vaccinated participants took the vaccination individually; therefore they could be vaccinated at different time points within May and December, and with different types of vaccines; most of them were vaccinated with the Pfizer vaccine (65%, 30 out of 46). Also, we aimed to select groups with the highest possible similarity of demographic parameters, either when compared between groups or when compared to the overall parameters of the whole Polish population. However, availability of participants who could meet these requirements was limited. A comparison of the demographics of the study groups with each other and with the general Polish population is presented in the supplementary material (Supplementary Figure S1 and S2).

### Patients hospitalized due to COVID-19

Anti-SARS-CoV-2 antibody levels were also analysed in non-vaccinated patients hospitalised for COVID-19 (over 18 years old, N=47) in the specialized COVID-19 hospital in Boleslawiec (Poland). Blood samples were collected at specific time points after the estimated infection, i.e. at day 5, at day 10, at day 15, at day 30, and at day 90. Depending on the course of the disease, patients were assigned to 3 groups: mild (N=21, no assisted respiratory therapy required), moderate (N=13, transient, non-invasive assisted respiratory therapy required/a mask), and severe/critical (N=12, assisted respiratory therapy with the nasal high-flow cannula or intensive non-invasive assisted respiratory therapy - BiPAP/CPAP or invasive mechanical ventilation). A comparison of the demographics of this group with the general Polish population is presented in the supplementary material (Supplementary Figure S3).

### Blood samples

Blood samples were collected in test tubes (BD SST II Advance), left to clot for 1 hour at room temperature (RT), and separated from the clot by centrifugation (15 min, 2000 g, RT) and then stored at – 20°C for further use.

### Serological diagnostic tests

Microblot-Array COVID-19 IgG and Microblot-Array COVID-19 IgA (TestLine Clinical Diagnostics s.r.o); cat.no. CoVGMA96, LOT 0100060496 and cat. no. CoVAMA96, LOT 0100066601); the arrays contain selected parts of the specific antigens NCP and RBD of SARS-CoV-2 virus.

### Identification of SARS-CoV-2 variants

In selected hospitalized patients, SARS-CoV-2 variants were identified by the targeted sequencing approach with the Ion AmpliSeq SARS-CoV-2 Insight Research Assay (Thermo Fisher Scientific, Europe, according to the manufacturer’s instructions). Briefly, patients were sampled by nasopharyngeal swabbing, viral RNA was isolated on silica spin columns (QIAquick) and the viral load was quantified by the qPCR standard diagnostic test. Ct values were used to design library preparation parameters, which included standard retrotranscription. Library preparation and sequencing chip loading were completed on Ion Chef. Sequencing was completed on Ion GeneStudio S5, and analysed with the software provided by the manufacturer. Sequences of sufficient quality were uploaded to the public database GISAID (https://www.gisaid.org/).

### Bioethics statements

The study was conducted in accordance with the principles of the Declaration of Helsinki. The research was approved by the local Bioethical Commission of the Regional Specialist Hospital in Wroclaw (approval number: KB/02/2020, policy No. COR193657). During the individual interview, all information about the study was provided and written consent was obtained from each participant. The written consent was accepted by the local Bioethical Commission of the Regional Specialist Hospital in Wroclaw (approval number: KB/02/2020).

### Statistical methods

Categorical variables are presented as absolute numbers and percentages in relation to the population under investigation. For quantitative variables, arithmetic means, first quartile (Q25%), median, third quartile (Q75%) and range of variability (extremes) were calculated. To determine the significance of differences between experimental subgroups, a one-way analysis of variance (ANOVA) and the Welsh test were performed. To assess differences in antibody levels between time points the t-test was used. Simple linear or logistic regression was used to assess the effect of selected variables on antibody levels and vaccination decisions. Chi^2^ test, Fisher’s test and ordinal logistic regression were used to calculate the relationship between obesity, anti-RBD/anti-NCP index and severity of the disease. Ordinal logistic regression was calculated using the MASS package in R programming language [14,15]. When appropriate, p-values were adjusted using the false discovery rate (FDR) method [16]. Statistical analyses were performed by GraphPad Prism 9.

## 3. Results

### Dynamics of anti-SARS-CoV-2 seroconversion in Polish population between May and December 2021

Four groups of participants were investigated: (1) (N-, V-) seronegative for SARS-CoV-2 antibodies in May 2021, non-vaccinated until December 2021, (2) (N-, V+) seronegative for SARS-CoV-2 antibodies in May 2021 but fully vaccinated between May and December 2021, (3) (N+, V-) seropositive for SARS-CoV-2-specific IgG antibodies in May 2021 but non-vaccinated until December 2021, (4) (N+, V+) seropositive for SARS-CoV-2-specific IgG antibodies in May 2021 and vaccinated between May and December 2021. Blood sera from these participants were used to determine levels of antibodies targeting SARS-CoV-2: NCP-specific or RBD-specific IgG, both qualitative (positive/negative) and quantitative (units per millilitre, U/ml) analysis was conducted.

In group 1 (N-, V-) we found 29 % (11 out of 38) participants positive for SARS-CoV-2-specific IgG (Table 1). This highlights the rate of unregistered SARS-CoV-2 infections that occurred between May and December in these participants. Twenty-six percent (10 out of 38) efficiently developed anti-RBD IgG, the most important antibody fraction in protection from COVID-19 due to their neutralizing potential [11]. However, the median increase of IgG levels in this group was very weak, consistent with the fact that the majority of the group had no effective contact with SARS-CoV-2 (Table 1, Table S1, Figure 1).

**Table 1.** Individuals identified as positive for anti-SARS-CoV-2 antibodies in the population without registered SARS-CoV-2 infections. The number of positive individuals and respective fraction (%) are presented. Antibodies were identified in blood samples after separation of blood sera by clotting and centrifugation. Microblot-Array testing was applied to qualify IgG level as positive or negative according to the cut-off given by the manufacturer, NCP – nucleocapsid protein, RBD – receptor binding protein.

|  |  | Group 1<br>(N=38)<br>negative for<br>anti-NCP IgG<br>in May.<br>non-vaccinated | Group 2<br>(N=29)<br>negative for<br>anti-NCP IgG<br>in May.<br>vaccinated | Group 3<br>(N=25)<br>positive for anti-NCP IgG<br>in May.<br>non-vaccinated | Group 4<br>(N=17)<br>positive for anti-NCP IgG<br>in May.<br>vaccinated |
| --- | --- | --- | --- | --- | --- |
| IgG | total positive | 11 (29%) | 27 (93%) | 19 (76%) | 17 (100%) |
|  | anti-NCP | 10 (26%) | 2 (7%) | 11 (44%) | 3 (18%) |
|  | anti-RBD | 10 (26%) | 27 (93%) | 13 (52%) | 17 (100%) |
| IgA | total positive | 8 (21%) | 16 (55%) | 13 (52%) | 17 (100%) |
|  | anti-NCP | 3 (8%) | 0 (0%) | 0 (0%) | 2 (12%) |
|  | anti-RBD | 7 (18%) | 15 (52%) | 12 (48%) | 17 (100%) |

**Figure 1.**
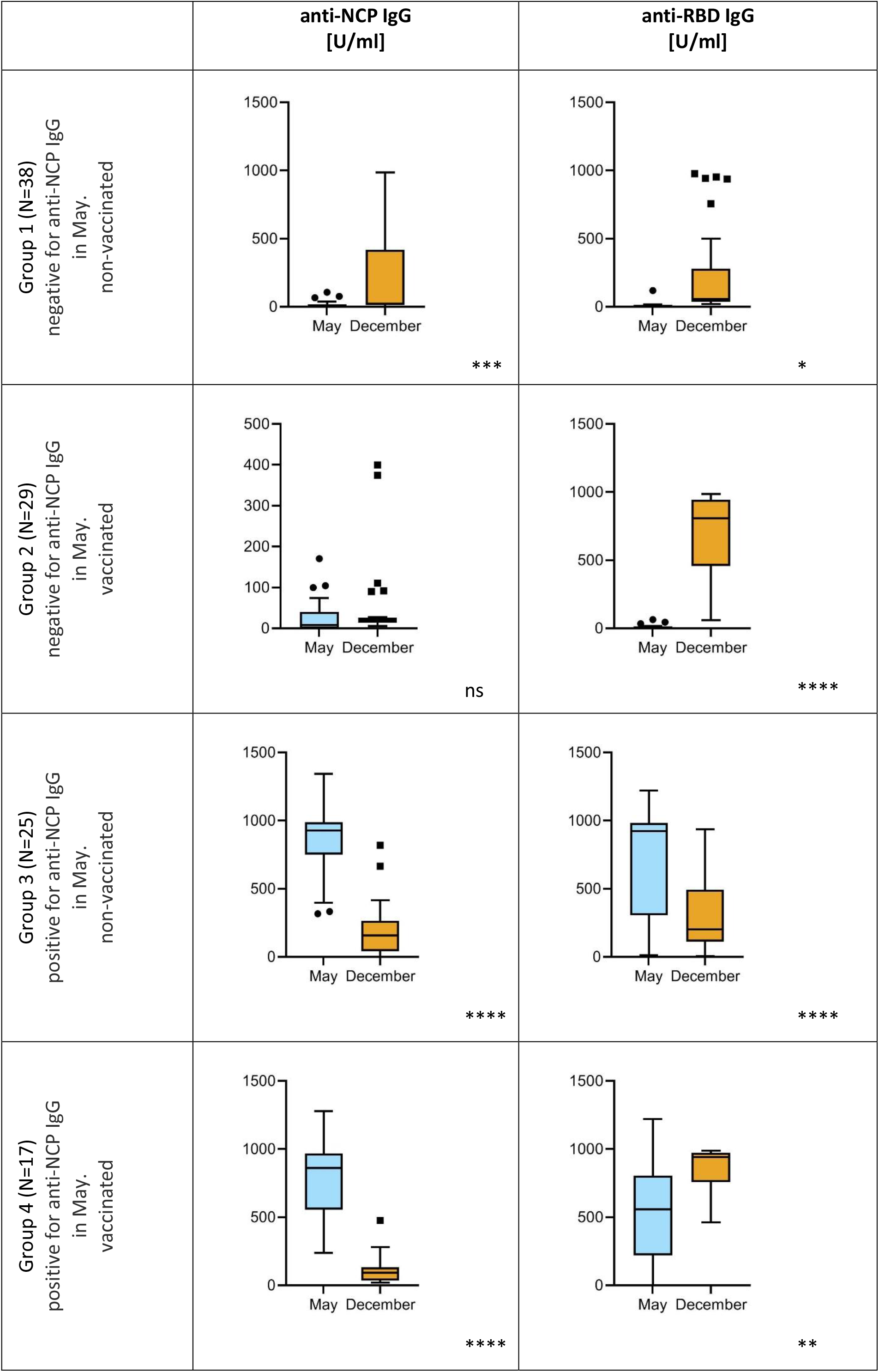
Comparison of SARS-CoV-2-specific IgG between May and December 2021 in population without registered SARS-CoV-2 infections. Antibodies were identified in blood samples after separation of blood sera by clotting and centrifugation. Microblot-Array testing was applied to quantify IgG level (U/ml), NCP – nucleocapsid protein, RBD – receptor binding protein. One-way analysis of variance (ANOVA) showed month to be a statistically significant factor (p<0.05) and Welsh’s t-test was used for comparisons between May and December results: ns – p>0.05; * – p<0.05; ** – p<0.01; *** – p<0.001; **** – p<0.0001

In group 2 (N-, V+), 93% (27 out of 29) of participants were found positive for SARS-CoV-2-specific IgG in December, with the same fraction of individuals who developed anti-RBD IgG (Table 1). This included a spectacular increase of the anti-RBD IgG quantitative median (from 0.398 to 809.6 U/ml) (Figure 1, Table S1), demonstrating the good efficacy of vaccination. Of note, vaccination provides anti-RBD immunization only, since antigens representing NCP are not included. Anti-NCP IgG can be developed in people who have been infected by SARS-CoV-2. Comparison of anti-NCP IgG in group 1 (non-vaccinated) and in group 2 (vaccinated) reveals that markedly more individuals were infected with SARS-CoV-2 between May and December in the first group (26% and 7%, respectively) (Table 1).

In group 3 (N+, V-), 76% (19 out of 25) of participants remained positive for any fractions of SARS-CoV-2-specific IgG also in December, and 13 of them (52%) presented detectable anti-RBD IgG (Table 1). However, the median value of this potentially virus neutralizing IgG fraction markedly decreased from 922.5 U/ml in May to 202.7 U/ml in December (which was very close to the negative cut-off value of the assay: 200 U/ml) (Figure 1, Table S1), thus demonstrating that the potential of SARS-CoV-2-specific IgG developed before May in protection against repeated infection at the end of the year was very limited (if any).

In group 4 (N+V+) 100% (17 out of 17) of participants tested positive for anti-SARS-CoV-2 IgG (Table 1). In this group also the highest levels of RBD-specific IgG were observed, both as the median value and as a maximum individual value achieved in any group (Figure 1, Table S1).

Overall levels of SARS-CoV-2 specific IgA revealed a similar tendency as IgG in all groups, though in most cases fewer individuals were positive for the investigated antibody fractions than those positive for IgG (Table 1).

### Seroconversion in patients hospitalized due to COVID-19

Seroconversion was further investigated in 47 unvaccinated patients hospitalized for COVID-19, since in this group time after infection could be estimated with a relatively high accuracy, thus making a reference for the general population (unregistered infections). Patients were categorized into three groups: (1) those showing a disease of mild severity, (2) those with a disease of moderate severity, and (3) those with a severe to critical course of the disease.

We did not find any significant differences in the antibody responses between infected male and female patients (Supplementary Figure S4, Supplementary Table S2); thus further analyses were conducted without gender grouping. We analysed the IgG kinetics at five different time points: day 5, day 10, day 15, day 30, and day 90 after infection estimated from symptoms onset.

From day 10, we observed an increase in both anti-RBD IgG and anti-NCP IgG antibodies, which reached high levels on post-infection day 30 and remained at high levels on post-infection day 90 (Figure 2). Available literature indicates an earlier and more intense immune response in patients with a severe course of coronavirus disease [10]; here the median values of both types of antibodies were higher in the group of patients with a severe course of the disease, but without statistical significance (p>0.05).

**Figure 2.**
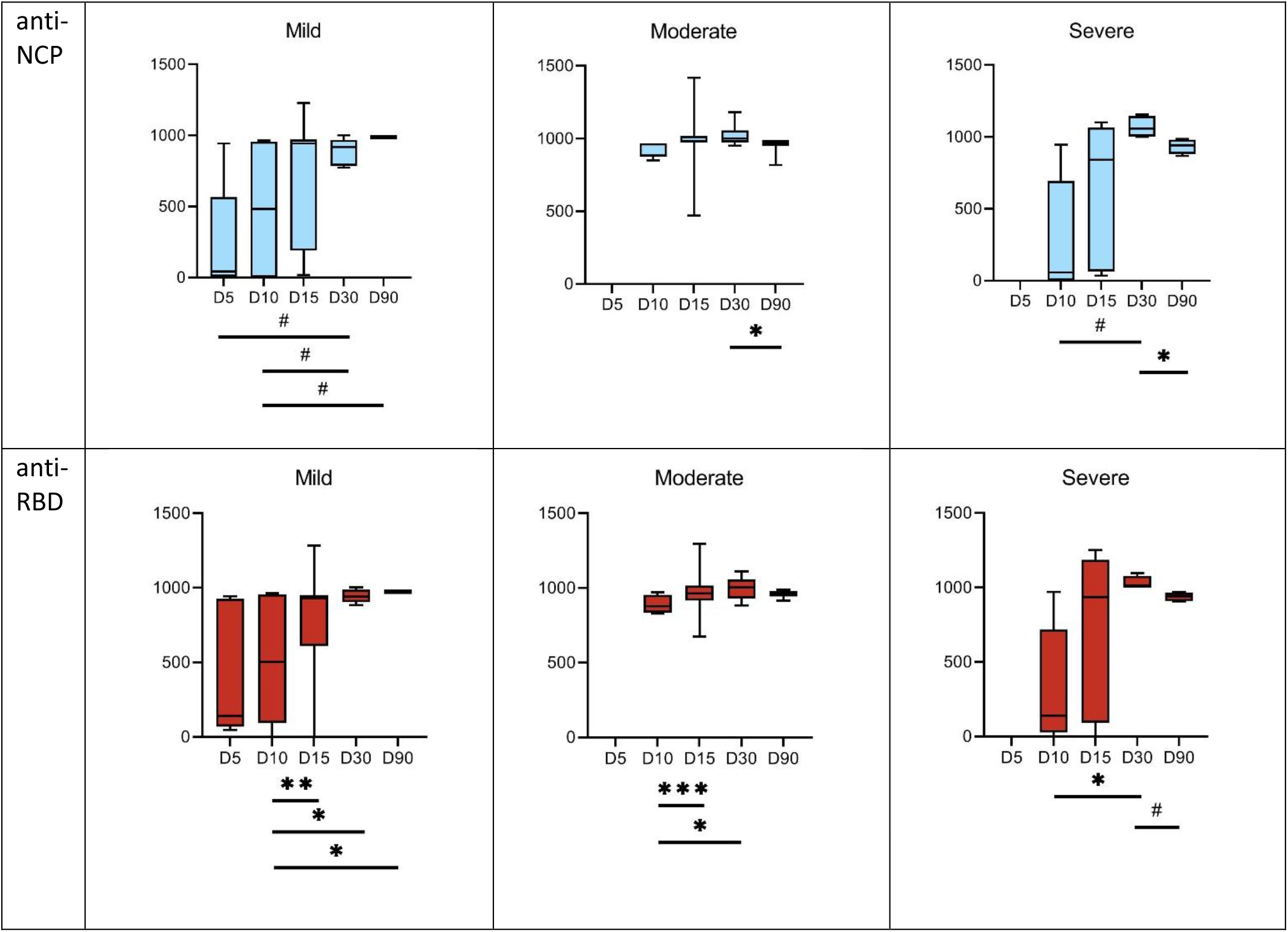
Kinetics of induction of SARS-CoV-2-specific IgG in hospitalized COVID-19 patients; anti-NCP IgG-fraction of IgG specific to nucleocapsid protein of SARS-CoV-2, anti-RBD IgG-fraction of IgG specific to receptor binding protein of SARS-CoV-2, mild – patients treated without assisted respiratory therapy, moderate – patients with limited support of non-invasive assisted respiratory therapy (mask), severe/critical – patients with intensive non-invasive assisted respiratory therapy or invasive mechanical ventilation, days – estimated number of days after onset of infection. Antibodies were identified in blood samples after separation of blood sera by clotting and centrifugation. Microblot-Array testing was applied to quantify IgG level (U/ml). T-test between each time point sample for anti-NCP and anti-RBD IgG titres was applied, # – 0.1 < p < 0.5; * – p<0.05; ** – p<0.01; *** – p<0.001

Interestingly, by analysis of the anti-N/anti-RBD IgG ratio in each patient at all time points, we found that at day 15 and 30 after infection, a high ratio correlated positively with chance of a mild course of the disease (p<0.05). Our observation in the hospitalized individuals is in line with literature reports presenting similar trends in asymptomatic and light COVID-19 cases [17]. Also, obesity correlated positively with the disease severity (p=0.0163), which increased the risk of moderate or severe course of the disease (p=0.005). Study reports confirm that obesity increases the risk of both COVID-19-related hospitalisations and death [18].

## Discussion

Among the structural proteins of the SARS-CoV-2 virus, spike protein containing the receptor binding domain (RBD) and nucleocapsid protein (NCP) have been found the most immunogenic, thus being first-choice targets for SARS-CoV-2 diagnostics. Evaluation of antibodies targeting RBD has also the highest potential for assessing possible immunological protection against SARS-CoV-2 infection, due to the neutralising potential of this antibody fraction. NCP-specific antibodies, in turn, are efficient markers of previous exposure to the virus and infection, and they are typically developed in parallel to those targeting RBD [9,19,20]. Of note, vaccination (with mRNA or adenoviral vector-based preparations available in Poland) provides anti-RBD immunization only, since antigens representing NCP are not included in the preparations. Therefore anti-NCP IgG can be developed as a result of SARS-CoV-2 virus infection but not vaccination, giving a clear indication that the infection occurred.

In this study we found a considerable fraction (26%) of anti-NCP IgG positive individuals in the population that had never been diagnosed for COVID-19 either before May 2021 or between May and December 2021. This reflects the considerable ‘dark figure’ of unregistered or even undiagnosed SARS-CoV-2 infections that however affect herd immunity. The same individuals (thus also 26%) demonstrated positive levels of anti-RBD IgG induced by the infection. Interestingly, the ‘dark figure’ from the previous pandemic waves, which was assessed in patients representing the same geographical region in May 2021, was highly comparable, at 25.6% [7]. Since pandemic waves before May 2021 in Poland were caused by the Alpha variant of the virus, while between May and December 2021 the Delta variant dominated, our observation indicates that the ‘dark figure’ has not changed with the SARS-CoV-2 variant. Of note, in group 2 (vaccinated, no previous infection), anti-NCP IgG was only observed in 2 individuals (7%), thus showing the efficacy of vaccination also in controlling unregistered or undiagnosed (probably asymptomatic to mild) infections with SARS-CoV-2 (Table 1).

As expected, high efficacy of vaccination for the induction of RBD-specific antibodies was revealed in both vaccinated groups (Table 1), although in 2 individuals (both vaccinated with adenoviral vector vaccines in August or September) no anti-SARS-CoV-2 antibodies were detected in December 2021. The literature so far confirms a decrease in the humoral response after vaccination over time, and studies comparing available vaccine types showed a considerable decrease at 8 months of follow-up for the J&J/Janssen vaccine [21,22]. Recommendations of the European Medicines Agency (EMA) and the European Centre for Disease Prevention and Control (ECDC) based on the analysis of available scientific evidence report higher immunogenicity and the associated increased efficacy of SARS-CoV-2 vaccines when a heterologous vectored/mRNA vaccine regimen is used, in comparison to immunization with a homologous adenoviral vector vaccines [23,24]. This may suggest that in our participants the level of protective antibodies fell to an undetectable level. Considering the relatively short time after vaccination (3-4 months), they also may belong to non-responders, that is, patients who do not develop antibodies after vaccination [25]. Interestingly, one of these patients provided information that she developed COVID-19 2 weeks after the study conducted in December.

The overall level of anti-RBD IgG was highest in the individuals who previously (before May 2021) were infected with SARS-CoV-2, and then they decided to take vaccination. All these participants (100%) were positive for anti-RBD IgG (Table 1), also demonstrating the highest levels of these antibodies, markedly higher than those in the individuals also infected with SARS-CoV-2 before May but not vaccinated (groups 3 and 4 in Figure 1). This observation highlights the positive role of vaccination in maintaining antiviral immunological protection in convalescents, and it is consistent with other studies. Trougakos et al. (2021) reported higher antibody titres induced by vaccination in those who recovered from COVID-19 than in healthy individuals [10]. Here we observed the same effect in individuals affected by asymptomatic to mild (and unregistered) infections.

This follow-up study was extended with an analysis of whether information that was given to participants on their serological status in May (n=501) affected their further decisions on vaccination. No correlation was found between levels of anti-NCP or anti-RBD antibodies in May and vaccination/non-vaccination later (data not shown), thus indicating that serological information given to participants had nether an encouraging nor a discouraging effect on them.

Due to the reports confirming the relationship between obesity, smoking, and lower antibody titres following COVID-19 vaccination [26], we performed analyses that showed no influence of BMI, smoking, or allergies on the level of produced antibodies in the studied groups (simple linear or logistic regression, Welsh test) (Supplementary Tables S3 and S4). However, the lack of correlation may be affected by the relatively small size of available subpopulations that were investigated.

We further analysed kinetics of antibody induction in patients hospitalized due to COVID-19. Of note, Alpha and Delta SARS-CoV-2 variants were identified in selected individuals, but no significant differences were found when comparing between variants (data not shown); thus we eventually presented summary results for all these patients. In hospitalized patients estimation of time of infection could be much better, and a clear increase of RBD- and NCP-specific antibodies within 2-4 weeks after infection was demonstrated (Figure 2). This observation is consistent with the study by Yamayoshi et al. (2021) where IgG titres against RBD proteins S and N peaked around 20 days after the onset of the disease, then gradually decreased and lasted for several months after the disease onset [27]. This observation sheds a light on the potential time that could have passed between asymptomatic or oligosymptomatic infections that resulted in seroconversion in unregistered convalescents. This time was probably between 2 weeks and a few months from the infection.

This observation suggests that antibodies observed in the general population (unregistered cases) could result mainly from the very recent, fourth pandemic wave in Poland, which started in mid-October and peaked around 28th November to 10th December, with 7-day averages exceeding 23 000 confirmed SARS-CoV-2 infections [13], thus making approximately 605 cases per one million citizens. This study further demonstrates that epidemiological analysis based on detection of anti-SARS-CoV-2 seroconversion allows for relatively rapid detection of recent cases, and it has a limited window of a few months after the pandemic peak of infections.

## Conclusions

In this study we continued population screening for hidden herd immunity to COVID-19 initiated in May 2021 [7], and we found that specific antibodies targeting SARS-CoV-2 that developed before May 2021 cannot be considered important 7 months later, in December 2021. The only factor that provided a good level of population immunity to COVID-19 was vaccination. The highest levels of protection were however achieved in the group that had been infected with SARS-CoV-2 before May 2021 and also fully vaccinated between May and December. This effect should be considered expected, since in that case the infection probably plays the role of an additional dose of viral antigens that boost the immune response. The so-called ‘dark figure’ of unregistered cases of SARS-CoV-2 infections remains relatively high at 26%, with an evident contribution of the very recent, fourth wave of the pandemic that occurred in Poland shortly before the study in December.

## Data Availability

All data produced in the present study are available upon reasonable request to the authors

## Acknowledgments

This work was supported by The National Centre for Research and Development in Poland, grant no. SZPITALEJEDNOIMIENNE/48/2020.

The authors are deeply grateful to all participants who agreed to act as serum donors in this study. Thank you for your help, support, understanding, and for your desire to contribute to scientific solutions to combat the disease.

## References

1. Kenneth M. Hirsh M. Bloom A. COVID-19: Epidemiology, virology, and prevention. UpToDate. https://www.uptodate.com/contents/covid-19-epidemiology-virology-and-prevention. Last updated Dec 14.2021.

2. Singh AK, Singh A, Singh R, Misra A. Molnupiravir in COVID-19: A systematic review of literature. Diabetes Metab Syndr. 2021 Nov-Dec;15(6):102329. doi: 10.1016/j.dsx.2021.102329. Epub 2021 Oct 30. PMID: 34742052; PMCID: PMC8556684.

3. Case JB, Winkler ES, Errico JM, Diamond MS. On the road to ending the COVID-19 pandemic: Are we there yet? Virology. 2021 May;557:70–85. doi: 10.1016/j.virol.2021.02.003. Epub 2021 Feb 26. PMID: 33676349; PMCID: PMC7908885.

4. Soeorg H, Jõgi P, Naaber P, Ottas A, Toompere K, Lutsar I. Seroprevalence and levels of IgG antibodies after COVID-19 infection or vaccination. Infect Dis (Lond). 2022 Jan;54(1):63–71. doi: 10.1080/23744235.2021.1974540. Epub 2021 Sep 14. PMID: 34520315; PMCID: PMC8442755.

5. Safiabadi Tali SH, LeBlanc JJ, Sadiq Z, Oyewunmi OD, Camargo C, Nikpour B, Armanfard N, Sagan SM, Jahanshahi-Anbuhi S. Tools and Techniques for Severe Acute Respiratory Syndrome Coronavirus 2 (SARS-CoV-2)/COVID-19 Detection. Clin Microbiol Rev. 2021 May 12;34(3):e00228–20. doi: 10.1128/CMR.00228-20. PMID: 33980687; PMCID: PMC8142517.

6. Oran DP, Topol EJ. Prevalence of Asymptomatic SARS-CoV-2 Infection : A Narrative Review. Ann Intern Med. 2020 Sep 1;173(5):362–367. doi: 10.7326/M20-3012. Epub 2020 Jun 3. PMID: 32491919; PMCID: PMC7281624.

7. Budziar W, Gembara K, Harhala M, Szymczak A, Jędruchniewicz N, Baniecki K, Pikies A, Nahorecki A, Hoffmann A, Kardaś A, Szewczyk-Dąbrowska A, Klimek T, Kaźmierczak Z, Witkiewicz W, Barczyk K, Dąbrowska K. Hidden fraction of Polish population immune to SARS-CoV-2 in May 2021. PLoS One. 2022 Feb 3;17(2):e0253638. doi: 10.1371/journal.pone.0253638. PMID: 35113873; PMCID: PMC8812878.

8. Bläckberg A, Fernström N, Sarbrant E, Rasmussen M, Sunnerhagen T. Antibody kinetics and clinical course of COVID-19 a prospective observational study. PLoS One. 2021 Mar 22;16(3):e0248918. doi: 10.1371/journal.pone.0248918. PMID: 33750984; PMCID: PMC7984607.

9. Szymczak A. Jędruchniewicz N. Torelli A. Kaczmarzyk-Radka A. Coluccio R. Kłak M. Konieczny A. Ferenc S. Witkiewicz W. Montomoli E. Miernikiewicz P. Bąchor R. Dąbrowska K. Antibodies specific to SARS-CoV-2 proteins N. S and E in COVID-19 patients in the normal population and in historical samples. J Gen Virol. 2021 Nov;102(11). doi: 10.1099/jgv.0.001692. PMID: 34816794.

10. Trougakos IP, Terpos E, Zirou C, Sklirou AD, Apostolakou F, Gumeni S, Charitaki I, Papanagnou ED, Bagratuni T, Liacos CI, Scorilas A, Korompoki E, Papassotiriou I, Kastritis E, Dimopoulos MA. Comparative kinetics of SARS-CoV-2 anti-spike protein RBD IgGs and neutralizing antibodies in convalescent and naïve recipients of the BNT162b2 mRNA vaccine versus COVID-19 patients. BMC Med. 2021 Aug 23;19(1):208. doi: 10.1186/s12916-021-02090-6. PMID: 34420521; PMCID: PMC8380479.

11. Tretyn A, Szczepanek J, Skorupa M, Jarkiewicz-Tretyn J, Sandomierz D, Dejewska J, Ciechanowska K, Jarkiewicz-Tretyn A, Koper W, Pałgan K. Differences in the Concentration of Anti-SARS-CoV-2 IgG Antibodies Post-COVID-19 Recovery or Post-Vaccination. Cells. 2021 Jul 31;10(8):1952. doi: 10.3390/cells10081952. PMID: 34440721; PMCID: PMC8391384.

12. Zhu L, Xu X, Zhu B, Guo X, Xu K, Song C, Fu J, Yu H, Kong X, Peng J, Huang H, Zou X, Ding Y, Bao C, Zhu F, Hu Z, Wu M, Shen H. Kinetics of SARS-CoV-2 Specific and Neutralizing Antibodies over Seven Months after Symptom Onset in COVID-19 Patients. Microbiol Spectr. 2021 Oct 31;9(2):e0059021. doi: 10.1128/Spectrum.00590-21. Epub 2021 Sep 22. PMID: 34550000; PMCID: PMC8557935.

13. Główny Urząd Statystyczny, 2022. Avaible: https://stat.gov.pl/covid/opracowania-covid-19/. [Data uzyskania dostępu: 20 02 2022]

14. Venables WN, Ripley BD (2002). Modern Applied Statistics with S, Fourth edition. Springer, New York. ISBN 0-387-954570.

15. R Core Team (2021). R: A language and environment for statistical computing. R Foundation for Statistical Computing, Vienna, Austria. https://www.R-project.org/

16. Benjamini, Y., and Hochberg, Y. (1995). Controlling the false discovery rate: a practical and powerful approach to multiple testing. Journal of the Royal Statistical Society Series B,57, 289–300. doi: 10.1111/j.2517-6161.

17. Tutukina M, Kaznadzey A, Kireeva M, Mazo I. IgG Antibodies Develop to Spike but Not to the Nucleocapsid Viral Protein in Many Asymptomatic and Light COVID-19 Cases. Viruses. 2021 Sep 28;13(10):1945. doi: 10.3390/v13101945. PMID: 34696374; PMCID: PMC8539461.

18. Huang Y, Lu Y, Huang YM, Wang M, Ling W, Sui Y, Zhao HL. Obesity in patients with COVID-19: a systematic review and meta-analysis. Metabolism. 2020 Dec;113:154378. doi: 10.1016/j.metabol.2020.154378. Epub 2020 Sep 28. PMID: 33002478; PMCID: PMC7521361.

19. Lo Sasso B. Giglio RV. Vidali M. Scazzone C. Bivona G. Gambino CM. Ciaccio AM. Agnello L. Ciaccio M. Evaluation of Anti-SARS-Cov-2 S-RBD IgG Antibodies after COVID-19 mRNA BNT162b2 Vaccine. Diagnostics (Basel). 2021 Jun 22;11(7):1135. doi: 10.3390/diagnostics11071135. PMID: 34206567; PMCID: PMC8306884.

20. Satarker S. Nampoothiri M. Structural Proteins in Severe Acute Respiratory Syndrome Coronavirus-2. Arch Med Res. 2020 Aug;51(6):482–491. doi: 10.1016/j.arcmed.2020.05.012. Epub 2020 May 25. PMID: 32493627; PMCID: PMC7247499.

21. Levin EG. Lustig Y. Cohen C. Fluss R. Indenbaum V. Amit S. Doolman R. Asraf K. Mendelson E. Ziv A. Rubin C. Freedman L. Kreiss Y. Regev-Yochay G. Waning Immune Humoral Response to BNT162b2 Covid-19 Vaccine over 6 Months. N Engl J Med. 2021 Dec 9;385(24):e84. doi: 10.1056/NEJMoa2114583. Epub 2021 Oct 6. PMID: 34614326; PMCID: PMC8522797.

22. Cohn BA. Cirillo PM. Murphy CC. Krigbaum NY. Wallace AW. SARS-CoV-2 vaccine protection and deaths among US veterans during 2021. Science. 2022 Jan 21;375(6578):331–336. doi: 10.1126/science.abm0620. Epub 2021 Nov 4. PMID: 34735261.

23. European Medicines Agency. Charakterystyka Produktu Leczniczego. Vaxzevria. [Online]. Available: http://www.ema.europa.eu. [Data uzyskania dostępu: 20 01 2022].

24. Urząd Rejestracji Produktów Leczniczych, Wyrobów Medycznych i Produktów Biobójczych. [Online]. Avaible: http://urpl.gov.pl/pl/rekomendacje-europejskiej-agencji-lek%C3%B3w-ema-i-europejskiego-centrum-ds-zapobiegania-i-kontroli. [Data uzyskania dostępu: 20 01 2022].

25. Polack FP, Thomas SJ, Kitchin N, Absalon J, Gurtman A, Lockhart S, Perez JL, Pérez Marc G, Moreira ED, Zerbini C, Bailey R, Swanson KA, Roychoudhury S, Koury K, Li P, Kalina WV, Cooper D, Frenck RW Jr, Hammitt LL, Türeci Ö, Nell H, Schaefer A, Ünal S, Tresnan DB, Mather S, Dormitzer PR, Şahin U, Jansen KU, Gruber WC; C4591001 Clinical Trial Group. Safety and Efficacy of the BNT162b2 mRNA Covid-19 Vaccine. N Engl J Med. 2020 Dec 31;383(27):2603–2615. doi: 10.1056/NEJMoa2034577. Epub 2020 Dec 10. PMID: 33301246; PMCID: PMC7745181.

26. Watanabe M, Balena A, Tuccinardi D, Tozzi R, Risi R, Masi D, Caputi A, Rossetti R, Spoltore ME, Filippi V, Gangitano E, Manfrini S, Mariani S, Lubrano C, Lenzi A, Mastroianni C, Gnessi L. Central obesity, smoking habit, and hypertension are associated with lower antibody titres in response to COVID-19 mRNA vaccine. Diabetes Metab Res Rev. 2022 Jan;38(1):e3465. doi: 10.1002/dmrr.3465. Epub 2021 May 11. PMID: 33955644; PMCID: PMC8209952.

27. Yamayoshi S, Yasuhara A, Ito M, Akasaka O, Nakamura M, Nakachi I, Koga M, Mitamura K, Yagi K, Maeda K, Kato H, Nojima M, Pattinson D, Ogura T, Baba R, Fujita K, Nagai H, Yamamoto S, Saito M, Adachi E, Ochi J, Hattori SI, Suzuki T, Miyazato Y, Chiba S, Okuda M, Murakami J, Hamabata T, Iwatsuki-Horimoto K, Nakajima H, Mitsuya H, Omagari N, Sugaya N, Yotsuyanagi H, Kawaoka Y. Antibody titers against SARS-CoV-2 decline, but do not disappear for several months. EClinicalMedicine. 2021 Feb;32:100734. doi: 10.1016/j.eclinm.2021.100734. Epub 2021 Feb 11. PMID: 33589882; PMCID: PMC7877219.

