## Supplementary material for "Dynamics of anti-SARS-CoV-2 seroconversion in individual patients and at the population level"

**Group 1**
anti-NCP IgG negative in May,
non-vaccinated till December

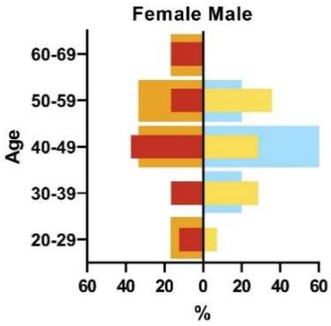

**Group 2**
anti-NCP IgG negative in May
vaccinated May-December

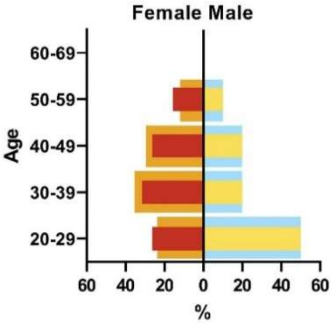

**Group 3**
anti-NCP IgG positive in May
non-vaccinated till December

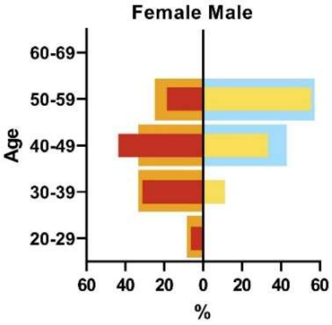

**Group 4**
anti-NCP IgG positive in May
vaccinated May-December

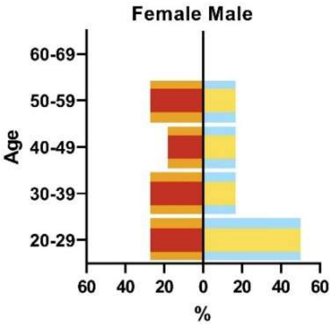

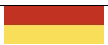

demographic parameters of each investigated group (backgraound)

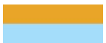

demographic parameters of the sub-population that was found immune to SARS-CoV-2 (IgG positive)

**Supplementary Figure S1.** Demographics in the serological screening of population without registered SARS-CoV-2 infections: demographic parameters of participants found positive for anti-SARS-CoV-2 IgG in each investigated group compared to general demographic parameters of the group. Antibodies were identified in blood samples after separation of blood sera by clotting and centrifugation. Microblot-array testing was applied to determine IgG level in each participants and positive/negative result was qualified according to the validated test.

<table>
<tr><td>Group 1<br>anti-NCP IgG negative in May,<br>non-vaccinated till December</td><td>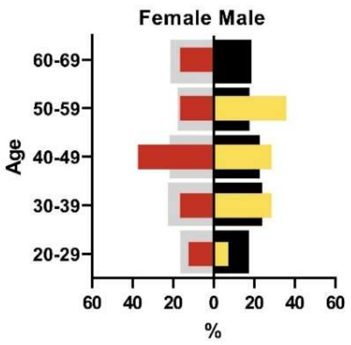
</td><td></td></tr>
<tr><td>Group 2<br>anti-NCP IgG negative in May<br>vaccinated May-December</td><td>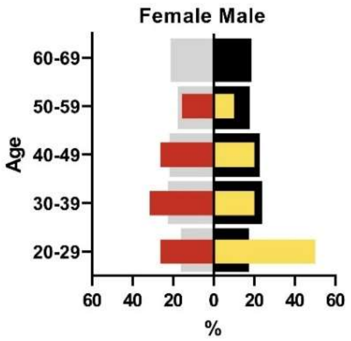
</td><td></td></tr>
<tr><td>Group 3<br>anti-NCP IgG positive in May<br>non-vaccinated till December</td><td>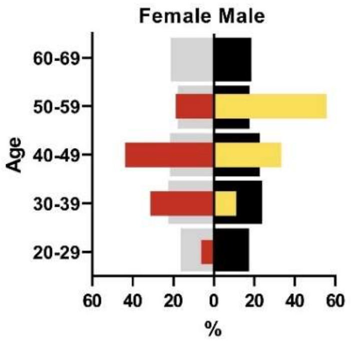
</td><td></td></tr>
<tr><td>Group 4<br>anti-NCP IgG positive in May<br>vaccinated May-December</td><td>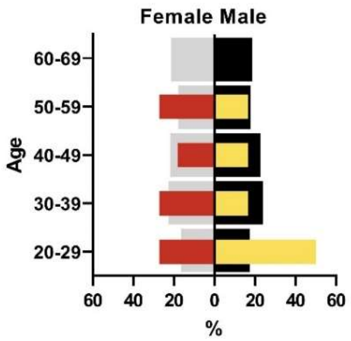
</td><td>demographic parameters of the Polish population (whole country, background)<br><br>demographic parameters of the investigated groups</td></tr>
</table>

**Supplementary Figure S2**. Demographics in the serological screening of population without registered SARS-CoV-2 infections: demographic parameters of each investigated group compared to general demographic parameters of the whole country.

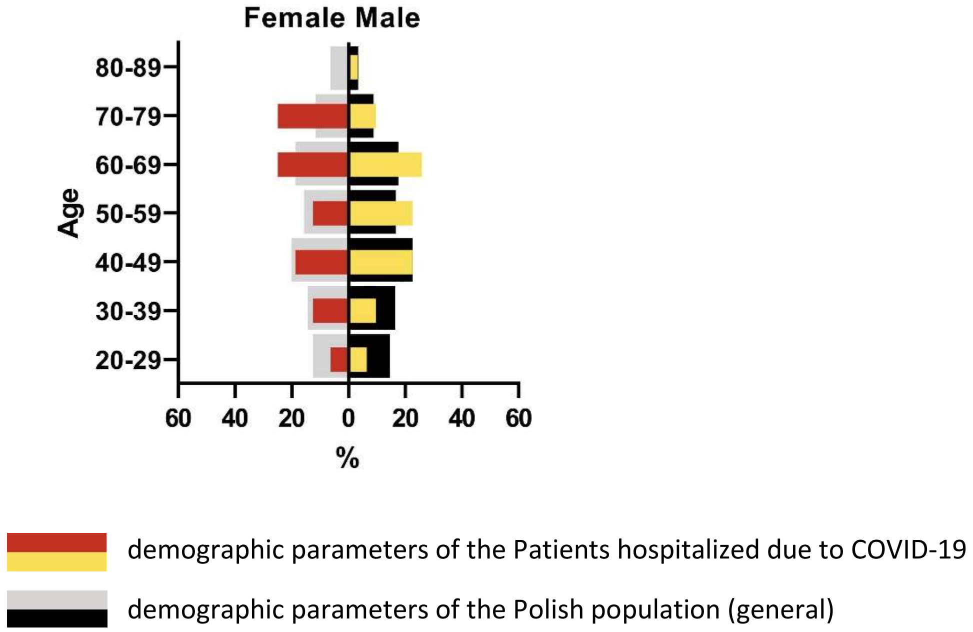

**Supplementary Figure S3.** Demographics in patients hospitalized due to COVID-19

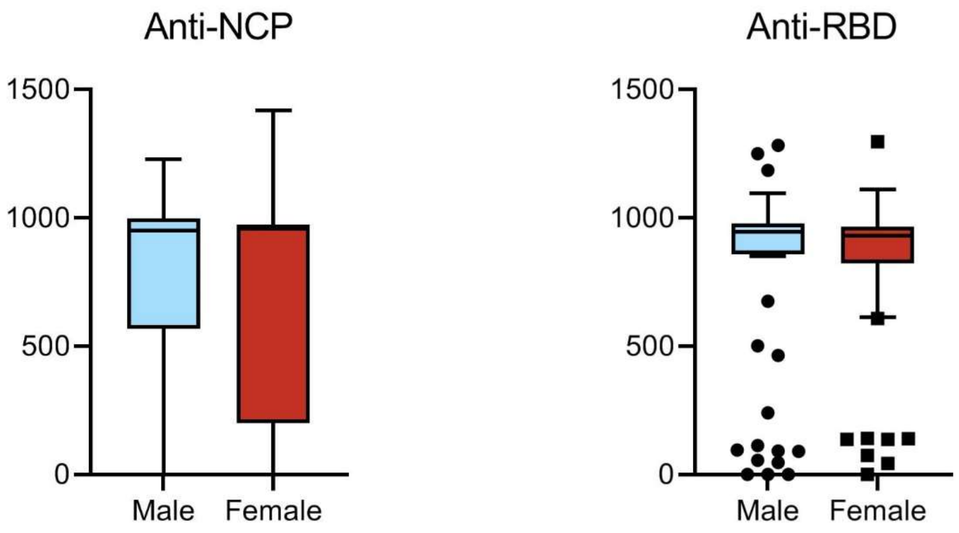

**Supplementary Figure S4**. IgG specific to SARS-CoV-2 in non-vaccinated patients hospitalized due to COVID-19: female and male comparison. Anti-NCP- antibodies specific to nucleocapsid protein N, anti-RBD- antibodies specific to receptor binding domain within spike protein S1 region. Antibodies were identified in blood samples after separation of blood sera by clotting and centrifugation. Microblot-array testing was applied to determine IgG level (U/ml). Median values are presented (dash), with quartiles 1 and 3 (boxes), and minimum/maximum values (whiskers); p>0.05 by Welsh test

| **Supplementary Table S1.** Distributions of quantitative variables of measurements for IgG specific to SARS-CoV-2 NCP and RBD in: Group 1: anti-NCP IgG negative, non-vaccinated; Group 2: anti-NCP IgG negative, vaccinated; Group 3: anti-NCP IgG positive, non-vaccinated; Group 4: anti-NCP IgG positive, vaccinated. NCP – nucleocapsid protein, RBD – receptor binding protein | | | | | | | | | | |
|---|---|---|---|---|---|---|---|---|---|---|
| **Group** | **Antibody** | **Date of the study** | **N** | **Min** | **Q25%** | **Median** | **Q75%** | **Max** | **Range** | **Average** |
| 1 | NCP | May | 38 | 0 | 0 | 5.211 | 17.27 | 107.3 | 107.3 | 13.11 |
| | | December | 38 | 0 | 13.82 | 20.96 | 416.5 | 986.2 | 986.2 | 219 |
| | RBD | May | 38 | 0 | 0 | 0.691 | 6.769 | 120 | 120 | 6.172 |
| | | December | 38 | 19.77 | 38.09 | 58.61 | 278.8 | 976.7 | 956.9 | 207.6 |
| 2 | NCP | May | 29 | 0 | 0.0605 | 8.425 | 39.6 | 170.7 | 170.7 | 27.53 |
| | | December | 29 | 5.701 | 14.55 | 21.6 | 26.16 | 400 | 394.3 | 51.78 |
| | RBD | May | 29 | 0 | 0 | 0.398 | 7.759 | 64.88 | 64.88 | 7.461 |
| | | December | 29 | 59.99 | 457.1 | 809.6 | 943.7 | 987.2 | 927.2 | 696.6 |
| 3 | NCP | May | 25 | 317 | 752 | 929 | 989.9 | 1343 | 1026 | 844.6 |
| | | December | 25 | 0 | 41.91 | 158.6 | 265.1 | 818.9 | 818.9 | 196.7 |
| | RBD | May | 25 | 11.73 | 305.2 | 922.5 | 982.9 | 1221 | 1209 | 683.2 |
| | | December | 25 | 5.202 | 112 | 202.7 | 492.1 | 935.5 | 930.3 | 308.9 |
| 4 | NCP | May | 17 | 238.1 | 553.9 | 862.1 | 969.6 | 1279 | 1041 | 774 |
| | | December | 17 | 19.61 | 35.1 | 91.53 | 133.6 | 476.9 | 457.2 | 118.8 |
| | RBD | May | 17 | 0 | 220.4 | 558.4 | 805.3 | 1221 | 1221 | 518.5 |
| | | December | 17 | 461.5 | 759 | 941.6 | 972.9 | 988.5 | 527 | 862 |
| Results are given as U/ml. Q25% and Q75% - 1. and 3. Quantile. | | | | | | | | | | |

| **Supplementary Table S2.** Distributions of quantitative variables – IgG specific to SARS-CoV-2 NCP, RBD and Spike S2 in: non-vaccinated patients hospitalized due to COVID-19 and vaccinated control group | | | | | | | | |
|---|---|---|---|---|---|---|---|---|
| | | N | Min | Q25% | Median | Q75% | Max | Average |
| non-vaccinated | NP | 95 | 0 | 469.3 | 957.1 | 988 | 1418 | 755.1 |
| | RBD | 95 | 0 | 830 | 940.4 | 972.5 | 1297 | 790.9 |
| male non-vaccinated | NP | 56 | 0 | 567 | 950.3 | 999.5 | 1229 | 776.3 |
| | RBD | 56 | 0 | 858.5 | 946 | 978.9 | 1283 | 797.1 |
| female non-vaccinated | NP | 40 | 0 | 200.9 | 957.7 | 974.5 | 1418 | 729.8 |
| | RBD | 40 | 0 | 823.2 | 932.1 | 965.2 | 1297 | 786.7 |
| Mild anti-NCP | D5 | 5 | 0 | 4.527 | 42.87 | 565.9 | 944.1 | 236.7 |
| | D10 | 11 | 0 | 3.067 | 483.3 | 957.2 | 964 | 474.6 |
| | D15 | 17 | 16.64 | 189.1 | 942.9 | 971.5 | 1229 | 657.8 |
| | D30 | 6 | 773.3 | 786 | 918.8 | 969.2 | 1002 | 893.5 |
| | D90 | 2 | 988 | 988 | 988.1 | 988.1 | 988.1 | 988.1 |
| Moderate anti-NCP | D10 | 4 | 850.4 | 877.3 | 961.3 | 967.7 | 968.7 | 935.4 |
| | D15 | 12 | 469.3 | 972.8 | 980.8 | 1018 | 1418 | 994.9 |
| | D30 | 8 | 950.2 | 974.2 | 1002 | 1057 | 1182 | 1022 |
| | D90 | 7 | 817.9 | 950.5 | 966.4 | 988.5 | 988.7 | 952.1 |

| | | | | | | | | |
|---|---|---|---|---|---|---|---|---|
| Severe anti-NCP | D10 | 5 | 0 | 0 | 58.18 | 693.2 | 945.1 | 288.9 |
| | D15 | 7 | 33.99 | 65.59 | 841.3 | 1066 | 1102 | 698.2 |
| | D30 | 4 | 1002 | 1004 | 1059 | 1145 | 1157 | 1069 |
| | D90 | 4 | 868.8 | 881.4 | 941.1 | 981.2 | 987.2 | 934.6 |
| Mild anti-RBD | D5 | 5 | 47.38 | 69.21 | 140.2 | 927.1 | 944.1 | 426.6 |
| | D10 | 11 | 0 | 96.13 | 501.9 | 957.2 | 964 | 516.9 |
| | D15 | 17 | 0 | 610.1 | 930.1 | 952.2 | 1283 | 771.3 |
| | D30 | 6 | 883.7 | 903.9 | 940.4 | 988.6 | 1004 | 943.8 |
| | D90 | 2 | 970.6 | 970.6 | 973.8 | 976.9 | 976.9 | 973.8 |
| Moderate anti-RBD | D10 | 4 | 830 | 835.1 | 878.3 | 954.1 | 970.1 | 889.2 |
| | D15 | 12 | 675.2 | 916.8 | 963.9 | 1015 | 1297 | 966.7 |
| | D30 | 8 | 882.9 | 932.2 | 1005 | 1056 | 1112 | 999.8 |
| | D90 | 7 | 915.3 | 944.6 | 959 | 977.6 | 988.7 | 957.9 |
| Severe Anti-RBD | D10 | 5 | 0 | 27.71 | 138.9 | 718.7 | 972.5 | 326.3 |
| | D15 | 7 | 0 | 92.36 | 936.4 | 1186 | 1250 | 760.6 |
| | D30 | 4 | 1002 | 1002 | 1013 | 1078 | 1096 | 1031 |
| | D90 | 4 | 906.8 | 909.3 | 940.2 | 967.3 | 968.6 | 938.9 |
| Results are given as U/ml. Q25% and Q75% - 1. and 3. Quantile. Day 5.10.15.30.90 from the onset of the infection. | | | | | | | | |

**Supplementary Table S3.** Number and percentage of smokers and allergies in studied groups.

| | Group 1 (N=38) negative for anti-NCP IgG in May. non-vaccinated | Group 2 (N=29) negative for anti-NCP IgG in May. vaccinated | Group 3 (N=25) positive for anti-NCP IgG in May. non-vaccinated | Group 4 (N=17) positive for anti-NCP IgG in May. vaccinated |
|---|---|---|---|---|
| Allergy | 11 (29%) | 7 (24%) | 4 (16%) | 6 (35%) |
| Smokers | 11 (29%) | 7 (24%) | 8 (32%) | 3 (18%) |

**Supplementary Table S4.** Distribution of BMI index in the study groups

| | Min | Q1 | Median | Q3 | Max | Mean | Std. Deviation | Std. Error of Mean |
|---|---|---|---|---|---|---|---|---|
| Group 1 (N=38) negative for anti-NCP IgG in May. non-vaccinated | 19.05 | 21.92 | 25.57 | 29.36 | 35.32 | 25.84 | 4.58 | 0.74 |
| Group 2 (N=29) negative for anti-NCP IgG in May. vaccinated | 17.1 | 21.81 | 23.83 | 28.36 | 33.79 | 24.79 | 4.56 | 0.85 |
| Group 3 (N=25) positive for anti-NCP IgG in May. non-vaccinated | 20.29 | 22.81 | 27.77 | 30.2 | 35.16 | 27.05 | 4.59 | 0.92 |

| | | | | | | | | |
|---|---|---|---|---|---|---|---|---|
| Group 4 (N=17) positive for anti-NCP IgG in May. vaccinated | 17.36 | 20.24 | 23.53 | 28.42 | 38.58 | 24.74 | 5.53 | 1.34 |
| Q25% and Q75% - 1. and 3. Quantile | | | | | | | | |

**Supplementary Table S5.** T-test between each time point sample for anti-RBD IgG titres. P-values obtained for each set of groups were adjusted for multiple hypothesis.

| Anti-RBD | | | | | |
|---|---|---|---|---|---|
| Mild | D5 | D10 | D15 | D30 | D90 |
| D5 | - | > 0.1 | > 0.1 | > 0.1 | > 0.1 |
| D10 | > 0.1 | - | **< 0.01** | **< 0.05** | **< 0.05** |
| D15 | > 0.1 | **< 0.01** | - | > 0.1 | > 0.1 |
| D30 | > 0.1 | **< 0.05** | > 0.1 | - | > 0.1 |
| D90 | > 0.1 | **< 0.05** | > 0.1 | > 0.1 | - |
| Moderate | D5 | D10 | D15 | D30 | D90 |
| D5 | - | - | - | - | - |
| D10 | - | - | **< 0.001** | **< 0.05** | > 0.1 |
| D15 | - | **< 0.001** | - | > 0.1 | > 0.1 |
| D30 | - | **< 0.05** | > 0.1 | - | > 0.1 |
| D90 | - | > 0.1 | > 0.1 | > 0.1 | - |
| Severe | D5 | D10 | D15 | D30 | D90 |
| D10 | > 0.1 | - | > 0.1 | **< 0.05** | > 0.1 |
| D15 | > 0.1 | > 0.1 | - | > 0.1 | > 0.1 |
| D30 | > 0.1 | **< 0.05** | > 0.1 | - | **0.1 > p > 0.05** |
| D90 | > 0.1 | > 0.1 | > 0.1 | **0.1 > p > 0.05** | - |

**Supplementary Table S6.** T-test between each time point sample for anti-NCP IgG titres.

| Anti -NCP | | | | | |
|---|---|---|---|---|---|
| Mild | D5 | D10 | D15 | D30 | D90 |
| D5 | - | > 0.1 | > 0.1 | **0.1 > p > 0.05** | > 0.1 |
| D10 | > 0.1 | - | > 0.1 | **0.1 > p > 0.05** | **0.1 > p > 0.05** |
| D15 | > 0.1 | > 0.1 | - | > 0.1 | > 0.1 |
| D30 | **0.1 > p > 0.05** | **0.1 > p > 0.05** | > 0.1 | - | > 0.1 |
| D90 | > 0.1 | **0.1 > p > 0.05** | > 0.1 | > 0.1 | - |
| Moderate | D5 | D10 | D15 | D30 | D90 |
| D5 | - | - | - | - | - |
| D10 | - | - | > 0.1 | > 0.1 | > 0.1 |

| | | | | | |
|---|---|---|---|---|---|
| D15 | - | > 0.1 | - | > 0.1 | > 0.1 |
| D30 | - | > 0.1 | > 0.1 | - | **< 0.05** |
| D90 | - | > 0.1 | > 0.1 | **< 0.05** | **-** |
| Severe | D5 | D10 | D15 | D30 | D90 |
| D10 | **0.1 > p > 0.05** | - | > 0.1 | **0.1 > p > 0.05** | > 0.1 |
| D15 | > 0.1 | > 0.1 | - | > 0.1 | > 0.1 |
| D30 | > 0.1 | **0.1 > p > 0.05** | > 0.1 | - | **< 0.05** |
| D90 | > 0.1 | > 0.1 | > 0.1 | **< 0.05** | - |
